# Response and role of palliative care during the COVID-19 pandemic: a national telephone survey of hospices in Italy

**DOI:** 10.1101/2020.03.18.20038448

**Authors:** Massimo Costantini, Katherine E Sleeman, Carlo Peruselli, Irene J Higginson

## Abstract

**Background:** Palliative care is an important component of healthcare in pandemics, contributing to symptom control, psychological support, and supporting triage and complex decision making.

**Aim:** To examine preparedness for, and impact of, the COVID-19 pandemic on hospices in Italy to inform the response in other countries.

**Design:** Cross-sectional telephone survey, carried out in March 2020.

**Setting:** Sixteen Italian hospices, purposively sampled according to COVID-19 risk into high (more than 25 COVID-19 cases per 100,000 inhabitants), medium (15-25 cases per 100,000), and low risk (fewer than 15 cases per 100,000) regions. A brief questionnaire was developed to guide the interviews. Descriptive analysis was undertaken.

**Results:** Seven high risk, five medium risk and four low risk hospices provided data. Two high risk hospices had experienced COVID-19 cases among both patients and staff. All hospices had implemented policy changes, and several had rapidly implemented changes in practice including transfer of staff from inpatient to community settings, change in admission criteria, and daily telephone support for families. Concerns included scarcity of personal protective equipment, a lack of hospice-specific guidance on COVID-19, anxiety about needing to care for children and other relatives, and poor integration of palliative care in the acute setting.

**Conclusion:** The hospice sector is capable of responding flexibly and rapidly to the COVID-19 pandemic. Governments must urgently recognise the essential contribution of hospice and palliative care to the COVID-19 pandemic, and ensure these services are integrated into the health care system response. Availability of personal protective equipment and setting-specific guidance is essential.

**What is already known:**

- The Coronavirus disease 2019 (COVID-19) has estimated global mortality of 3.4%, and numbers of cases are rapidly escalating worldwide.
- Hospice services face unprecedented pressure, with resources rapidly stretched beyond normal bounds.
- No data exist on the response and role of hospice and palliative care teams to COVID-19.
- Within Europe, Italy has been most affected by COVID-19.

**What this paper adds:**

- We surveyed 16 Italian hospices in March 2020, all of which had implemented rapid policy changes in response to COVID-19.
- Changes to practice included moving to more support in community settings, change in admission criteria, and daily telephone support for families.
- Personal protective equipment and guidance were lacking.
- Assessments of risk and potential impact on staff varied greatly.

**Implications for policy and practice:**

- Governments must recognise the hospice and palliative care sector as an essential component of the health care system response to COVID-19.
- The hospice sector is capable of responding rapidly to the COVID-19 pandemic, but the potential of this response will be undermined unless hospices can access personal protective equipment.
- Considerations for hospice services during the COVID-19 pandemic are changes to visitor policies, interruption of volunteering, shifting roles and responsibilities such as greater community working and telephone support for relatives.

## Introduction

Coronavirus disease 2019 (COVID-19), caused by severe acute respiratory syndrome coronavirus 2 (SARS-CoV-2), emerged in Wuhan, China, in December 2019. On 11^th^ March 2020 COVID-19 was declared a pandemic by the WHO. The WHO estimates global mortality at 3.4%,^1^ though mortality rates are higher among older people and those with comorbidities. A cohort of 36 non-survivors of COVID-19 in China identified the most prevalent symptoms as fever (94%), shortness of breath (58%), fatigue (47%) and cough (39%).^2^

Within Europe, Italy has been most affected by COVID-19. The first case was identified on 21^st^ February 2020, and as of 15^th^ March 2020 more than 24,000 cases and 1,809 deaths have been recorded in Italy. Escalating numbers of deaths are anticipated elsewhere. Palliative care is an essential component of healthcare in pandemics, contributing to symptom control, as well as psychological support for patients, carers and health care professionals, and supporting triage and complex decision making.^3^ However, hospices may be particularly vulnerable to disruption in pandemics, and very little data exists on the response of, or impact on, palliative care services in these situations.^4^ The aim of this study was to examine preparedness for, and impact of, COVID-19 on hospices in Italy in order to inform the response in other countries.

## Method

### Design

We conducted a telephone survey of 16 Italian hospices. We purposively sampled hospices from regions at different risk, according to the number of COVID-19 positive patients per 100,000 inhabitants: low risk (less than 15 cases × 100,000), medium risk (between 15 and 25 cases per 100,000) and high risk (more than 25 per 100,000).

### Procedures

Hospices were contacted by telephone by MC or CP between 11^th^ and 15^th^ March 2020, and an interview with the medical or nursing director was requested and arranged for a suitable time. Interviews took place by telephone and lasted between 10 and 15 minutes. A brief questionnaire was developed by the authors to guide the interviews (appendix 1), including the opportunity for free text comments (Appendix 1). A Likert scale was used to understand perceptions of risk (0=no risk, 10=maximum risk imaginable).

### Analysis

Descriptive analysis was undertaken of quantitative and qualitative data.

### Ethics

The ethics committee of Reggio Emilia was approached, and we were advised that according to Italian law a formal ethical approval was not necessary.

## Results

All hospices approached provided data. Their size ranged from 7 to 34 beds (mean 15.9, SD 8.3) (Table 1). Four were in low risk areas, five were in medium risk areas, and seven were in high risk areas. Interviews were conducted with 13 medical directors and three directors of nursing. 11 hospices were public, and five were private (not for profit). Seven hospices were affiliated with acute hospitals, including two hospitals that had been designated exclusively COVID-19 positive hospitals. Two of the ‘high risk’ hospices had experienced cases of COVID-19 positivity. Cases included both patients and staff (including nurses, physicians and health care assistants).

**Table 1:** Characteristics of the hospices that provided data and their rating of effects on workforce and risk. * This unit includes 8 beds that are affiliated to a hospital ** Scale was 0=no risk, 10=maximum risk imaginable

| Hospice ID | Area | Hospice characteristics |  |  | COVID-19 positive cases | Effects on workforce (Likert scale 0-10)** |  | Perception of risk (Likert scale 0-10)** |  |
| --- | --- | --- | --- | --- | --- | --- | --- | --- | --- |
|  |  | Number of beds | Hospice type | Hospital association |  | Anxiety about needing to care for children | Anxiety about needing to care for relatives | Of staff becoming unwell | Of hospice closure |
| 1 | Low risk | 10 | Public | Yes | No | 5 | 5 | 2 | 1 |
| 2 |  | 25 | Private non-profit | No | No | 8 | 3 | 0 | 0 |
| 3 |  | 7 | Public | No | No | 7 | 8 | 5 | 2 |
| 4 |  | 10 | Public | Yes | No | 6 | 6 | 3 | 0 |
| 5 | Medium risk | 11 | Public | No | No | 6 | 8 | 4 | 0 |
| 6 |  | 12 | Public | Yes | No | 3 | 3 | 5 | 3 |
| 7 |  | 34 | Private non-profit | No | No | 6 | 3 | 6 | 2 |
| 8 |  | 10 | Public | Yes | No | 5 | 4 | 9 | 5 |
| 9 |  | 29 | Public | Yes* | No | 10 | 10 | 7 | 0 |
| 10 | High risk | 30 | Private non-profit | No | No | 8 | 8 | 6 | 3 |
| 11 |  | 10 | Public | Yes | Yes | 7 | 7 | 3 | 0 |
| 12 |  | 16 | Public | No | No | 6 | 6 | 5 | 4 |
| 13 |  | 16 | Private non-profit | No | Yes | 1 | 3 | 2 | 2 |
| 14 |  | 12 | Private non-profit | No | No | 6 | 6 | 8 | 7 |
| 15 |  | 10 | Public | Yes | No | 7 | 7 | 1 | 0 |
| 16 |  | 12 | Public | No | No | 3 | 3 | 3 | 1 |

### Procedures and guidance

Most hospices agreed that there was written guidance covering procedures in the event that patients, relatives or staff members either tested positive for COVID-19 or were suspected cases. All hospices followed national and regional guidance, in six cases guidelines of the local trust. None used guidelines or procedures specific for hospices though in one case guidance was described as locally defined but ‘strengthened’ for the hospice. No hospices had written guidance that referred specifically to volunteers, and 15/16 stated that they were no longer using volunteers

### Personal protective equipment

Equipment including gloves, masks and disposable gowns were being used, though use and provision varied considerably. One physician in a high risk area noted use of *“very rigorous dressing procedures including FP2 masks and special overalls”* while in another high risk area the interviewee commented *“no mandatory protection, professionals can choose what to do”*. A third interviewee (high risk area) noted use of *“gloves and masks… but there is a great scarcity of this equipment”*.

### Changes in visiting policies

All hospices had changed visitor policies, though there was not a unified approach to this. 12 had adopted a policy of allowing only one relative per patient. Two of these hospices (high risk areas) were willing to relax this policy when patients where dying, while one (medium risk area) only allowed visitors when patients were dying. One hospice (high risk area) required that visitors needed to remain in the hospice day and night and that they could not return once they had left the hospice, and two (high and medium risk areas) had completely closed to visitors. One hospice (medium risk area) comprised two separate units, one of which had adopted a ‘one visitor only’ policy, and the other had closed to visitors. Two hospices (medium risk areas) were screening relatives for symptoms before entering the hospice. Of the limited visiting hours, one interviewee (high risk area) noted *“relatives seem to understand what the staff are doing and appreciate their work”*.

### Admission criteria

Most hospices reported no change in their admission criteria, though one had cancelled respite admissions and closed to admissions from hospitals. Three hospices had implemented a telephone triage system before admission to assess risk of COVID-19 positivity. Two hospices (high risk areas) were openly accepting patients infected with COVID-19, who were being isolated in specific areas of the hospices. One hospice (high risk area) had a policy not to admit patients with COVID-19.

### Care after death

Care after death varied, with four hospices limiting the number of relatives who could view the body of the deceased patient. One hospice (high risk area) had banned any relatives from entering the mortuary, and another (high risk area) had adopted a system where relatives viewed the body of the deceased through a window.

### Impact on workforce

Hospices reported moderate levels of staff anxiety about the need to care for either children (mean response 5.4 on Likert scale 0-10) or other relatives (mean response 5.7), and there was little difference between high, medium and low risk areas. Several hospices reported that staff were worried about coming to work, for example one nurse coordinator (medium risk hospice) reported *“staff are very worried and agitated about the risk to themselves and the possibility of taking the virus home”*. Nevertheless, staff absence was low, with one physician commenting *“the staff are scared but still they go to work”*.

### Assessment of risk

Hospices perceived a moderate risk of hospice staff being infected with COVID-19 over the coming week (mean response 4.0 on Likert scale 0-10). This was higher in medium (6.2) and high (4.0) risk areas than in low risk areas (2.5). Hospices in low risk areas perceived a low risk of the hospice closing in the coming week because of infected staff members (mean response 0.75 on Likert scale). This was greater in medium (2.0) and high risk (2.4) areas though remained low.

### Changes in practice

Several hospices had rapidly implemented changes in practice. One hospice (high risk area) had noted a reduction in requests for admission so had moved staff from inpatient to home care services. Another (high risk area), where visiting had been severely limited, had implemented a system where the hospice psychologist was telephoning patients’ relatives every day to update them and provide psychological support. Other hospices had cancelled all internal meetings, as well as annual leave.

### Psychological impact on staff

A lack of adequate preparation made caring for COVID-19 positive patients difficult: one medical director noted *“Positive patients entered and we were not prepared”*. Several interviewees spoke of the difficulty of providing holistic care within the constraints of an infectious disease outbreak *“It is difficult to maintain the humanity of palliative care in this situation”*. This included the acute setting *“Guidance on care for people dying from COVID-19 is missing”*, while one physician noted *“People with this infection are dying in ICU very badly, without any kind of palliative care support”*. The impact of COVID-19 on care of the dying was felt to reach beyond the acute illness *“At the end of this story, I think palliative care in Italy and everywhere will be very different from before”*.

## Discussion

### Main findings

We provide the first data from the hospice sector of preparedness for, and impact of, COVID-19. At the time of data collection, two hospices (both in high risk areas) were aware of having COVID-19 positive patients or staff. However, all had implemented changes in policies in response to COVID-19, for example concerning visitors and volunteers, and several had rapidly implemented changes in practice according to changing needs.

An important concern voiced by staff was a lack of preparedness for COVID-19. While all hospices in our survey had written guidance on procedures for suspected and confirmed cases of COVID-19, this was mostly regionally or nationally provided and no hospice had setting-specific guidance. There was considerable variation in the use of barrier precautions and personal protective equipment, which were described as scarce. There was also wide variation in perceptions of anxiety and the risk of illness among staff. This may indicate that more and urgent education is needed to inform hospice staff about reducing risks of COVID-19 infection. Protection of health care professionals across all settings against COVID-19 through use of appropriate barrier precautions should be of the highest priority, to avoid illness and mitigate against psychological distress.^1^

Provision of holistic care in the context of an infectious disease outbreak was noted to be highly challenging. Hospices responded to this challenge through rapid changes to service provision. For example one hospice had implemented daily telephone calls to relatives who were unable to visit, which might mitigate against the ‘disruption in connectedness’ described following the 2003 SARS epidemic in Singapore.^5^ Another hospice had moved staff from inpatient to home care services as a result of a falling number of inpatient referrals. Changes in inpatient hospice utilisation in Taiwan were identified during and after the SARS epidemic, and led to recommendations to distribute hospice care services into networks (e.g. home care, acute hospital inpatient care and inpatient hospice care) that can adapt to changing needs.^4^

### Strengths and limitations

The limitations of this study are that it was a rapid telephone survey with a small sample. The opportunity to collect in depth qualitative data was limited due to the extreme pressure services are under. It will be useful to follow up this cross-sectional survey over time as the situation changes. Using our existing clinical-academic networks enabled this survey to be completed rapidly, to provide essential early data of the hospice response.

### What this study adds

The hospice sector has an important role to play in the response to COVID-19. This includes support with complex decisions and triage, psychological support for patients, carers and professionals, and complex symptom management, particularly for people who are dying.^3^ However, there is evidence that the hospice sector is underused in epidemics.^4^ In Italy, and elsewhere, it is likely that the number of people dying with COVID-19 will overwhelm the capacity of the acute sector.^6^ Integrating hospice and palliative care into acute services may free up resources to optimise survival of others.^7^

Our data highlights that hospice services in all countries need to act now to prepare for COVID-19. Building on the Critical Care model of providing surge capacity in a crisis, elements essential to implementing a palliative care pandemic plan include (i) medication and equipment for symptom control including kits for use in care homes and at home; (ii) education to frontline staff on symptom management and end-of-life care including developing standardized order sheets and protocols, and involving allied care workers in providing psychological and bereavement support; (iii) identification of wards and beds appropriate to accommodate patients expected to die; and (iv) systems to identify patients in need of palliative care and to provide appropriate support across settings.^8^

## Conclusions

Hospices are uniquely placed to rapidly develop expertise in holistic care for people with COVID-19, including direct care of the dying as well as facilitating advance care planning in anticipation of acute deterioration. Our survey demonstrates that the hospice sector is able to respond flexibly and rapidly to the COVID-19 pandemic. However, the potential of hospices in supporting the COVID-19 pandemic will be undermined unless the sector has access to appropriate protective equipment and setting-specific guidance. Governments must urgently recognise the necessity of hospice and palliative care to the COVID-19 pandemic, ensure these services are both protected and integrated into the health care system response.

## Data Availability

Data is available from the corresponding author

## Contributions

MC, CP conceived the idea, MC, CP developed the protocol with input from IJH. MC, CP conducted the interviews. KES led analysis of the data with critical input from all authors. IJH conducted the search for evidence with input from all authors. KES with MC wrote the first draft of the paper. All authors contributed to critical review.

## References

1 Adams JG, Walls RM. Supporting the Health Care Workforce During the COVID-19 Global Epidemic. JAMA. 2020 Mar 12. doi: 10.1001/jama.2020.3972.

2 Huang Y, Yang R, Xu Y, Gong P. Clinical characteristics of 36 non-survivors with COVID-19 in Wuhan, China. Preprint available at https://www.medrxiv.org/content/10.1101/2020.02.27.20029009v2

3 Nouvet E, Sivaram M, Bezanson K et al. Palliative care in humanitarian crises: a review of the literature. Journal of International Humanitarian Action. 2018 3:5. doi.org/10.1186/s41018-018-0033-8

4 Chen TJ, Lin MH, Chou LF, Hwang SJ. Hospice utilization during the SARS outbreak in Taiwan. BMC Health Serv Res. 2006 Aug 4;6:94. doi:10.1186/1472-6963-6-94

5 Leong IY, Lee AO, Ng TW, et al. The challenge of providing holistic care in a viral epidemic: opportunities for palliative care. Palliat Med. 2004 Jan;18(1):12–8.

6 Remuzzi A, Remuzzi G. COVID-19 and Italy: what next? Lancet. 2020 March 12. doi.org/10.1016/S0140-6736(20)30627-9

7 Matzo M, Wilkinson A, Lynn J, Gatto M, Phillips S. Palliative Care Considerations in Mass-Casualty Events with Scarce Resources. Biosecur Bioterror. 2009 Jun;7(2):199–210. doi: 10.1089/bsp.2009.0017.

8 Downar J, Seccareccia D. Palliating a pandemic: “all patients must be cared for”. J Pain Symptom Manage. 2010 Feb;39(2):291–5. doi:10.1016/j.jpainsymman.2009.11.241.

